# Characterization and clinical course of 1000 patients with COVID-19 in New York: retrospective case series

**DOI:** 10.1101/2020.04.20.20072116

**Authors:** Michael G Argenziano, Samuel L Bruce, Cody L Slater, Jonathan R Tiao, Matthew R Baldwin, R Graham Barr, Bernard P Chang, Katherine H Chau, Justin J Choi, Nicholas Gavin, Parag Goyal, Angela M Mills, Ashmi A Patel, Marie-Laure S Romney, Monika M. Safford, Neil W Schluger, Soumitra Sengupta, Magdalena E Sobieszczyk, Jason E Zucker, Paul A Asadourian, Fletcher M Bell, Rebekah Boyd, Matthew F Cohen, MacAlistair I Colquhoun, Lucy A Colville, Joseph H de Jonge, Lyle B Dershowitz, Shirin A Dey, Katherine A Eiseman, Zachary P Girvin, Daniella T Goni, Amro A Harb, Nicholas Herzik, Sarah Householder, Lara E Karaaslan, Heather Lee, Evan Lieberman, Andrew Ling, Ree Lu, Arthur Y Shou, Alexander C Sisti, Zachary E Snow, Colin P Sperring, Yuqing Xiong, Henry W Zhou, Karthik Natarajan, George Hripcsak, Ruijun Chen

**Affiliations:** Columbia University Vagelos College of Physicians and Surgeons, New York, NY; Department of Biomedical Informatics at Columbia University Medical Center, New York, NY; Division of Pulmonary, Allergy and Critical Care Medicine, Department of Medicine, NewYork-Presbyterian/Columbia University Irving Medical Center, New York, NY; Division of General Medicine, Department of Medicine, NewYork-Presbyterian/Columbia University Irving Medical Center, New York, NY; Department of Emergency Medicine, NewYork-Presbyterian/Columbia University Irving Medical Center, New York, NY; Division of Cardiology, Department of Medicine, NewYork-Presbyterian/Columbia University Irving Medical Center, New York, NY; Department of Medicine, Weill Cornell Medicine, New York, NY; Division of Infectious Diseases, Department of Medicine, NewYork-Presbyterian/Columbia University Irving Medical Center, New York, NY

**Author notes:** Mr. Argenziano, Mr. Bruce, Mr. Slater, and Mr. Tiao contributed equally to this work. Corresponding Authors: RuiJun Chen MD and George Hripcsak MD, RuiJun Chen MD, Department of Biomedical Informatics, Columbia University Irving Medical Center, 622 West 168th St., PH-20 New York, NY 10032.

## Abstract

**Objective:** To characterize patients with coronavirus disease 2019 (COVID-19) in a large New York City (NYC) medical center and describe their clinical course across the emergency department (ED), inpatient wards, and intensive care units (ICUs).

**Design:** Retrospective manual medical record review.

**Setting:** NewYork-Presbyterian/Columbia University Irving Medical Center (NYP/CUIMC), a quaternary care academic medical center in NYC.

**Participants:** The first 1000 consecutive patients with laboratory-confirmed COVID-19.

**Methods:** We identified the first 1000 consecutive patients with a positive RT-SARS-CoV-2 PCR test who first presented to the ED or were hospitalized at NYP/CUIMC between March 1 and April 5, 2020. Patient data was manually abstracted from the electronic medical record.

**Main outcome measures:** We describe patient characteristics including demographics, presenting symptoms, comorbidities on presentation, hospital course, time to intubation, complications, mortality, and disposition.

**Results:** Among the first 1000 patients, 150 were ED patients, 614 were admitted without requiring ICU-level care, and 236 were admitted or transferred to the ICU. The most common presenting symptoms were cough (73.2%), fever (72.8%), and dyspnea (63.1%). Hospitalized patients, and ICU patients in particular, most commonly had baseline comorbidities including of hypertension, diabetes, and obesity. ICU patients were older, predominantly male (66.9%), and long lengths of stay (median 23 days; IQR 12 to 32 days); 78.0% developed AKI and 35.2% required dialysis. Notably, for patients who required mechanical ventilation, only 4.4% were first intubated more than 14 days after symptom onset. Time to intubation from symptom onset had a bimodal distribution, with modes at 3-4 and 9 days. As of April 30, 90 patients remained hospitalized and 211 had died in the hospital.

**Conclusions:** Hospitalized patients with COVID-19 illness at this medical center faced significant morbidity and mortality, with high rates of AKI, dialysis, and a bimodal distribution in time to intubation from symptom onset.

## INTRODUCTION

Coronavirus disease 2019 (COVID-19) is a global pandemic, with New York City (NYC) as an epicenter of the disease. Since the first confirmed case of COVID-19 on March 1, 2020, there were 164,505 laboratory-confirmed cases across the city, resulting in 42,417 hospitalizations and 13,000 confirmed deaths (as of April 30).^1^ Internationally, the rapid spread of COVID-19 taxed hospital system resources and drove a scarcity of ventilators and other medical equipment in many countries.^2^ Within NYC, the high burden of disease quickly exceeded the standard capacity of hospital systems, requiring massive expansion of inpatient and intensive care unit (ICU) facilities and raising concerns regarding optimal clinical management, safe maximization of hospital throughput, and resource allocation.^3^ ^4^

Despite the pressing need for evidence to inform such key decisions, data remain limited on COVID-19 in the U.S., and how it compares with previously published international cohorts. Patient characteristics, illness course, practice patterns, resource utilization, morbidity, and mortality associated with COVID-19 have been characterized in only limited samples.^5–9^ The U.S. effort at characterizing this disease began with two small case series from Seattle while internationally, Wuhan, China^10–12^ and Lombardy, Italy^13^ have published more extensively about their experiences. Characteristics of NYC patients are beginning to be enumerated with limited data on hospitalized patients, including the critically ill,^14^ but it remains largely unknown how these patients compare to previously described U.S. and international cohorts and what implications these differences will have on clinical care, outcomes, and resources.^6^ ^15^

Therefore, we sought to characterize the course of the first 1000 consecutive adult COVID-19 patients at NewYork-Presbyterian/Columbia University Irving Medical Center (NYP/CUIMC), a large quaternary care academic medical center. We provide a detailed description of demographic data, comorbidities, presenting symptoms, clinical course including time to intubation, hospital complications, patient outcomes, and mortality. In Box 1, we additionally provide the overall clinical context driving care throughout the first months of the pandemic’s spread in NYC.

#### Box 1 Criteria for Testing and Treatment

##### Criteria for COVID-19 Testing and Diagnosis

Testing Policies
- Early March – Recommended testing only hospitalized, symptomatic patients
- Mid March – Updated to include patients exhibiting symptoms and who required hospitalization, were at high risk, or were being discharged to congregate settings.
- Early April – Expanded to all patients being admitted to the hospital.

Diagnosis
- A COVID-19 diagnosis was defined as a positive result on the RT-PCR assay for SARS-CoV-2.

##### Criteria for Hospital and ICU Admission

Hospital Admission
- Most common criteria for admission to hospital was room air hypoxemia.

ICU Admission
- Most commonly reserved for patients with acute respiratory failure requiring mechanical ventilation.

##### Criteria for Intubation and Extubation

Intubation
- Initiated for patients with hypoxemia on a non-rebreather face mask or high flow nasal cannula oxygen therapy (SpO_2_ 88–92%) and/or substantial increased work of breathing, altered mental status, or arterial hypotension.
- Self-proning was encouraged for patients requiring a non-rebreather face mask or high flow nasal cannula oxygen therapy who were alert and able to self-prone.
- Extubation was sought for patients who:

1. Had improving, mild hypoxemia (SpO_2_ > 90% with FiO_2_ ≤ 40%).
2. Passed a spontaneous breathing trial using pressure support ventilation.
3. Were hemodynamically stable.
4. Had a Richmond Agitation Scale Score^16^ ≥ -2

## METHODS

### Data source and study sample

We used data from the NYP/CUIMC electronic health record (EHR) and NYP clinical data warehouse to identify patients with laboratory-confirmed COVID-19 infection, as represented by a positive SARS-CoV-2-RT-PCR test. NYP/CUIMC Clinical Microbiology Laboratory began in-house testing on March 11th, with earlier tests sent out to the New York Department of Health (DOH), and the latest initial positive test for this cohort on April 6^th^. Patients in this cohort with positive DOH tests all had repeat positive tests at NYP/CUIMC. This aligned with patients who initially presented between March 1 to April 5, 2020. We performed ongoing retrospective manual data abstraction from EHR charts of all patients with COVID-19 who received ED or inpatient care at NYP/CUIMC (excluding tests performed in the outpatient setting or at an outside hospital). We characterized the first 1000 consecutive patients with COVID-19.

NYP/CUIMC is a quaternary care academic medical center with 765 adult beds serving a diverse, high acuity patient population in the Manhattan borough of NYC.^17^ NYP/CUIMC includes Milstein Hospital, which includes 6 ICUs, and Milstein Heart Center, which includes an additional coronary and cardiothoracic ICU, for a total of 117 adult ICU beds. As patient volume increased, an additional 160 surge ICU beds were created to expand capacity in multiple locations throughout the hospital. The non-ICU general medicine bed capacity was expanded from 216 to 540. With the increase in capacity and resources, all necessary treatments and interventions remained available to patients throughout the study period. For the purpose of this paper, an ICU bed was defined as one with the capability of providing mechanical ventilation and continuous vital sign monitoring, with staffing by critical care nurses and oversight by intensivists. The most common criterion for hospital admission for COVID-19 patients was room air hypoxemia. ICU admissions were mostly commonly reserved for patients with acute respiratory failure requiring mechanical ventilation.

### Manual chart review

Supervised by multiple clinicians and informaticians, an abstraction team of 30 trained medical students from the Columbia University Vagelos College of Physicians and Surgeons manually abstracted EHR data in chronological order by test date. Information from the charts was inputted directly into REDCap,^18^ using an instrument previously designed and validated by the abstraction team at Weill Cornell Medicine, who identified a mean Cohen’s kappa for categorical variables of 0.92 (IQR 0.86–0.97) and 0.94 (IQR 0.87–0.97) for continuous variables.^6^ The REDCap instrument collects 274 data fields, 90 of which are required. Our abstraction team was trained in multiple hour-long sessions by the Weill Cornell team and instrument developers. Calibration of the data collection across both sites was achieved through biweekly meetings and use of remote communication platforms. Records with missing data or with inconsistent times were reviewed by a second, dedicated quality control abstractor. A random subsample of abstracted data was checked by a second abstractor, typically a clinician, for calibration and consistency. Any conflicting data were resolved by consensus.

Data collected included demographics, comorbidities, presenting symptoms, laboratory and radiographic findings, hospital course including admission, ICU transfer, mechanical ventilation, complications (defined as those documented by clinicians in the EHR) such as acute respiratory distress syndrome (ARDS) or acute kidney injury (AKI), and disposition including discharge, transfer, or death. Supplemental Table 1 lists the definitions used to define the specified complications. Time of first symptom was recorded based on the patient’s history; if the patient did not or could not give a specific date of first symptom, it was recorded that the patient could only give an approximate time. Data that were not present in the EHR were excluded from analysis; no imputation was performed. Laboratory test data and race/ethnicity data was extracted from the clinical data warehouse.

### Data characterization and analysis

Individual records were labeled with the highest level of care a patient received as of April 30: (1) ED only, (2) hospitalized (non-ICU), and (3) ICU admission. This paper includes those still hospitalized, discharged, or who died in the hospital. For patients with multiple COVID-19 related visits recorded in the EHR, the visit with the highest level of care was selected. For patients with multiple visits with the same level of care, the most recent visit was selected. Characteristics were stratified by the highest level of care received to date, and 95% confidence intervals (CIs) were recorded for each value. A multivariate Cox proportional hazards analysis was performed to predict death, intubation, and a composite of either death or intubation.

All analyses and visualizations were performed using R.^19^ Continuous variables were reported as medians and interquartile ranges (IQR). Relevant time differences were computed from EHR documented dates and times. Hartigan’s dip test was used to test for multimodality.^20^

## Results

Between March 11^th^ and April 6^th^, a total of 2,423 patients were tested for SARS-CoV-2 at NYP/CUIMC, with 1,403 patients testing positive and 1,020 negative (Supplemental Table 2). Of the patients with a positive test, 1132 tested received ED or hospital care. Our cohort includes the first 1000 consecutive of these patients. Of this 1000 patient sample, there were 150 ED patients, 614 hospitalized patients who did not require ICU-level care, and 236 ICU patients; 90 patients were still hospitalized as of April 30.

### Baseline characteristics

Table 1 presents a detailed breakdown of baseline characteristics including demographics, comorbidities, and home medications. The median (IQR) age was 63.0 (50.0-75.0) years. There was a male predominance in the overall sample (59.6%) which was more pronounced among ICU patients (66.9% male). The median (IQR) body mass index (BMI) for all patients was 28.6 (25.2–33.1) kg/m^2^ and 29.4 (25.7 − 34.2) kg/m^2^ for ICU patients. Hypertension was the most common comorbidity, present in 60.1% of patients, followed by diabetes in 37.2% (Table 1). Only 8.2% of patients reported no major comorbidities. The most common home medications were statins (36.1%) and ACE-inhibitors (ACEi) or angiotensin receptor blockers (ARBs) (28.4%).

**Table 1.**
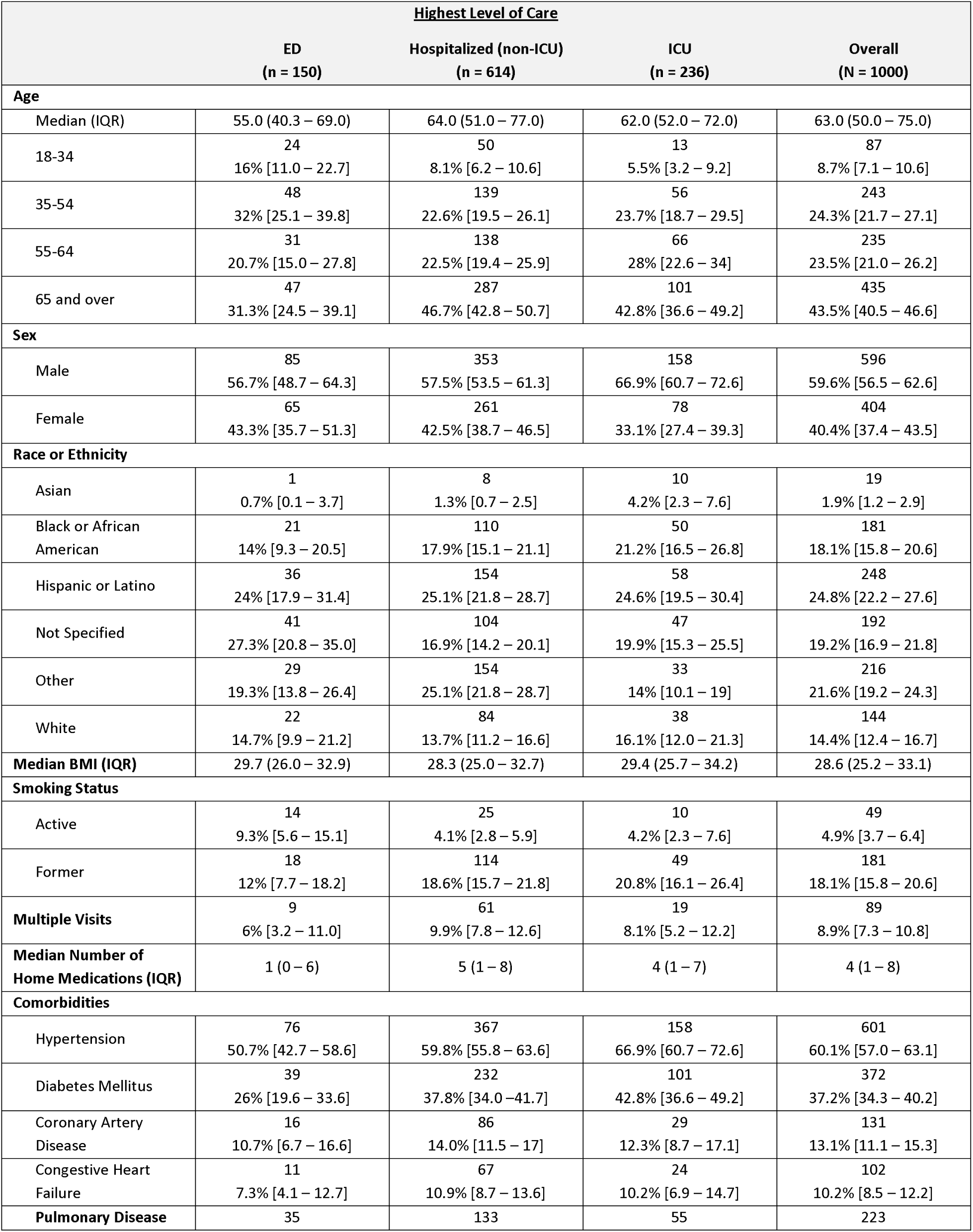
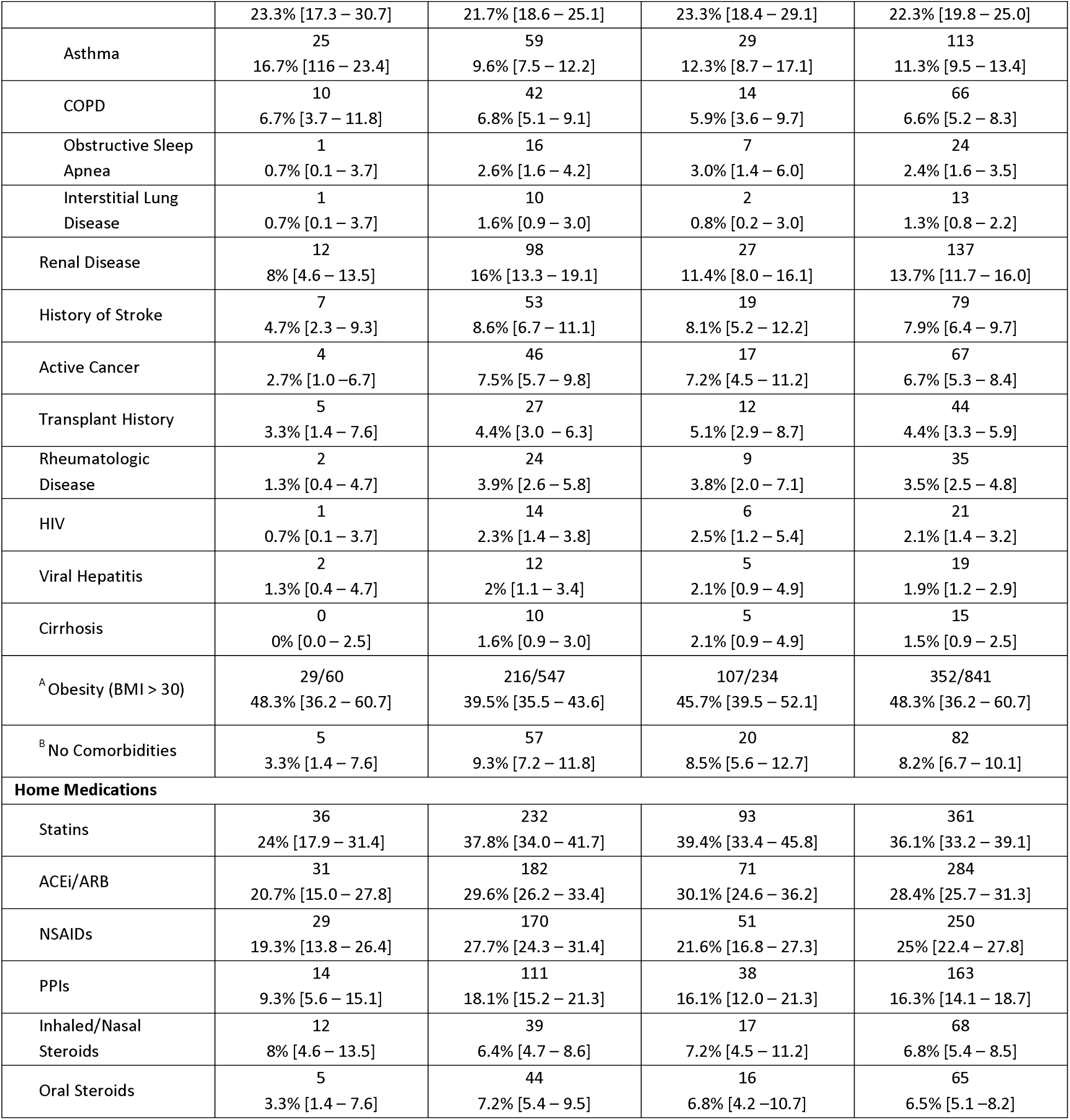
Baseline Characteristics. COVID-19 positive patients were stratified according to their highest level of care. For patients who sought care multiple times, their highest level of care is reported. All categorical data is reported as a frequency and a column percentage [95% confidence interval]. All continuous data is reported as median (interquartile range). ACEi, Angiotensin converting enzyme inhibitor; ARB, angiotensin receptor blocker; NSAIDs, nonsteroidal anti-inflammatory drugs; PPI, proton pump inhibitor. ^A^ For obesity denominators are reported due to incomplete reporting for BMI. ^B^ No Major Comorbidities indicates patients who have none of the listed comorbidities

| **Highest Level of Care** |  |  |  |  |
| --- | --- | --- | --- | --- |
|  | **ED**<br>**(n = 150)** | **Hospitalized (non-ICU)**<br>**(n = 614)** | **ICU**<br>**(n = 236)** | **Overall**<br>**(N = 1000)** |
| **Age** |  |  |  |  |
| Median (IQR) | 55.0 (40.3 – 69.0) | 64.0 (51.0 – 77.0) | 62.0 (52.0 – 72.0) | 63.0 (50.0 – 75.0) |
| 18-34 | 24<br>16% [11.0 – 22.7] | 50<br>8.1% [6.2 – 10.6] | 13<br>5.5% [3.2 – 9.2] | 87<br>8.7% [7.1 – 10.6] |
| 35-54 | 48<br>32% [25.1 – 39.8] | 139<br>22.6% [19.5 – 26.1] | 56<br>23.7% [18.7 – 29.5] | 243<br>24.3% [21.7 – 27.1] |
| 55-64 | 31<br>20.7% [15.0 – 27.8] | 138<br>22.5% [19.4 – 25.9] | 66<br>28% [22.6 – 34] | 235<br>23.5% [21.0 – 26.2] |
| 65 and over | 47<br>31.3% [24.5 – 39.1] | 287<br>46.7% [42.8 – 50.7] | 101<br>42.8% [36.6 – 49.2] | 435<br>43.5% [40.5 – 46.6] |
| **Sex** |  |  |  |  |
| Male | 85<br>56.7% [48.7 – 64.3] | 353<br>57.5% [53.5 – 61.3] | 158<br>66.9% [60.7 – 72.6] | 596<br>59.6% [56.5 – 62.6] |
| Female | 65<br>43.3% [35.7 – 51.3] | 261<br>42.5% [38.7 – 46.5] | 78<br>33.1% [27.4 – 39.3] | 404<br>40.4% [37.4 – 43.5] |
| **Race or Ethnicity** |  |  |  |  |
| Asian | 1<br>0.7% [0.1 – 3.7] | 8<br>1.3% [0.7 – 2.5] | 10<br>4.2% [2.3 – 7.6] | 19<br>1.9% [1.2 – 2.9] |
| Black or African American | 21<br>14% [9.3 – 20.5] | 110<br>17.9% [15.1 – 21.1] | 50<br>21.2% [16.5 – 26.8] | 181<br>18.1% [15.8 – 20.6] |
| Hispanic or Latino | 36<br>24% [17.9 – 31.4] | 154<br>25.1% [21.8 – 28.7] | 58<br>24.6% [19.5 – 30.4] | 248<br>24.8% [22.2 – 27.6] |
| Not Specified | 41<br>27.3% [20.8 – 35.0] | 104<br>16.9% [14.2 – 20.1] | 47<br>19.9% [15.3 – 25.5] | 192<br>19.2% [16.9 – 21.8] |
| Other | 29<br>19.3% [13.8 – 26.4] | 154<br>25.1% [21.8 – 28.7] | 33<br>14% [10.1 – 19] | 216<br>21.6% [19.2 – 24.3] |
| White | 22<br>14.7% [9.9 – 21.2] | 84<br>13.7% [11.2 – 16.6] | 38<br>16.1% [12.0 – 21.3] | 144<br>14.4% [12.4 – 16.7] |
| **Median BMI (IQR)** | 29.7 (26.0 – 32.9) | 28.3 (25.0 – 32.7) | 29.4 (25.7 – 34.2) | 28.6 (25.2 – 33.1) |
| **Smoking Status** |  |  |  |  |
| Active | 14<br>9.3% [5.6 – 15.1] | 25<br>4.1% [2.8 – 5.9] | 10<br>4.2% [2.3 – 7.6] | 49<br>4.9% [3.7 – 6.4] |
| Former | 18<br>12% [7.7 – 18.2] | 114<br>18.6% [15.7 – 21.8] | 49<br>20.8% [16.1 – 26.4] | 181<br>18.1% [15.8 – 20.6] |
| **Multiple Visits** | 9<br>6% [3.2 – 11.0] | 61<br>9.9% [7.8 – 12.6] | 19<br>8.1% [5.2 – 12.2] | 89<br>8.9% [7.3 – 10.8] |
| **Median Number of Home Medications (IQR)** | 1 (0 – 6) | 5 (1 – 8) | 4 (1 – 7) | 4 (1 – 8) |
| **Comorbidities** |  |  |  |  |
| Hypertension | 76<br>50.7% [42.7 – 58.6] | 367<br>59.8% [55.8 – 63.6] | 158<br>66.9% [60.7 – 72.6] | 601<br>60.1% [57.0 – 63.1] |
| Diabetes Mellitus | 39<br>26% [19.6 – 33.6] | 232<br>37.8% [34.0 – 41.7] | 101<br>42.8% [36.6 – 49.2] | 372<br>37.2% [34.3 – 40.2] |
| Coronary Artery Disease | 16<br>10.7% [6.7 – 16.6] | 86<br>14.0% [11.5 – 17] | 29<br>12.3% [8.7 – 17.1] | 131<br>13.1% [11.1 – 15.3] |
| Congestive Heart Failure | 11<br>7.3% [4.1 – 12.7] | 67<br>10.9% [8.7 – 13.6] | 24<br>10.2% [6.9 – 14.7] | 102<br>10.2% [8.5 – 12.2] |
| **Pulmonary Disease** | 35 | 133 | 55 | 223 |
|  | 23.3% [17.3 – 30.7] | 21.7% [18.6 – 25.1] | 23.3% [18.4 – 29.1] | 22.3% [19.8 – 25.0] |
| Asthma | 25<br>16.7% [11.6 – 23.4] | 59<br>9.6% [7.5 – 12.2] | 29<br>12.3% [8.7 – 17.1] | 113<br>11.3% [9.5 – 13.4] |
| COPD | 10<br>6.7% [3.7 – 11.8] | 42<br>6.8% [5.1 – 9.1] | 14<br>5.9% [3.6 – 9.7] | 66<br>6.6% [5.2 – 8.3] |
| Obstructive Sleep Apnea | 1<br>0.7% [0.1 – 3.7] | 16<br>2.6% [1.6 – 4.2] | 7<br>3.0% [1.4 – 6.0] | 24<br>2.4% [1.6 – 3.5] |
| Interstitial Lung Disease | 1<br>0.7% [0.1 – 3.7] | 10<br>1.6% [0.9 – 3.0] | 2<br>0.8% [0.2 – 3.0] | 13<br>1.3% [0.8 – 2.2] |
| Renal Disease | 12<br>8% [4.6 – 13.5] | 98<br>16% [13.3 – 19.1] | 27<br>11.4% [8.0 – 16.1] | 137<br>13.7% [11.7 – 16.0] |
| History of Stroke | 7<br>4.7% [2.3 – 9.3] | 53<br>8.6% [6.7 – 11.1] | 19<br>8.1% [5.2 – 12.2] | 79<br>7.9% [6.4 – 9.7] |
| Active Cancer | 4<br>2.7% [1.0 – 6.7] | 46<br>7.5% [5.7 – 9.8] | 17<br>7.2% [4.5 – 11.2] | 67<br>6.7% [5.3 – 8.4] |
| Transplant History | 5<br>3.3% [1.4 – 7.6] | 27<br>4.4% [3.0 – 6.3] | 12<br>5.1% [2.9 – 8.7] | 44<br>4.4% [3.3 – 5.9] |
| Rheumatologic Disease | 2<br>1.3% [0.4 – 4.7] | 24<br>3.9% [2.6 – 5.8] | 9<br>3.8% [2.0 – 7.1] | 35<br>3.5% [2.5 – 4.8] |
| HIV | 1<br>0.7% [0.1 – 3.7] | 14<br>2.3% [1.4 – 3.8] | 6<br>2.5% [1.2 – 5.4] | 21<br>2.1% [1.4 – 3.2] |
| Viral Hepatitis | 2<br>1.3% [0.4 – 4.7] | 12<br>2% [1.1 – 3.4] | 5<br>2.1% [0.9 – 4.9] | 19<br>1.9% [1.2 – 2.9] |
| Cirrhosis | 0<br>0% [0.0 – 2.5] | 10<br>1.6% [0.9 – 3.0] | 5<br>2.1% [0.9 – 4.9] | 15<br>1.5% [0.9 – 2.5] |
| <sup>A</sup> Obesity (BMI > 30) | 29/60<br>48.3% [36.2 – 60.7] | 216/547<br>39.5% [35.5 – 43.6] | 107/234<br>45.7% [39.5 – 52.1] | 352/841<br>48.3% [36.2 – 60.7] |
| <sup>B</sup> No Comorbidities | 5<br>3.3% [1.4 – 7.6] | 57<br>9.3% [7.2 – 11.8] | 20<br>8.5% [5.6 – 12.7] | 82<br>8.2% [6.7 – 10.1] |
| **Home Medications** |  |  |  |  |
| Statins | 36<br>24% [17.9 – 31.4] | 232<br>37.8% [34.0 – 41.7] | 93<br>39.4% [33.4 – 45.8] | 361<br>36.1% [33.2 – 39.1] |
| ACEi/ARB | 31<br>20.7% [15.0 – 27.8] | 182<br>29.6% [26.2 – 33.4] | 71<br>30.1% [24.6 – 36.2] | 284<br>28.4% [25.7 – 31.3] |
| NSAIDs | 29<br>19.3% [13.8 – 26.4] | 170<br>27.7% [24.3 – 31.4] | 51<br>21.6% [16.8 – 27.3] | 250<br>25% [22.4 – 27.8] |
| PPIs | 14<br>9.3% [5.6 – 15.1] | 111<br>18.1% [15.2 – 21.3] | 38<br>16.1% [12.0 – 21.3] | 163<br>16.3% [14.1 – 18.7] |
| Inhaled/Nasal Steroids | 12<br>8% [4.6 – 13.5] | 39<br>6.4% [4.7 – 8.6] | 17<br>7.2% [4.5 – 11.2] | 68<br>6.8% [5.4 – 8.5] |
| Oral Steroids | 5<br>3.3% [1.4 – 7.6] | 44<br>7.2% [5.4 – 9.5] | 16<br>6.8% [4.2 – 10.7] | 65<br>6.5% [5.1 – 8.2] |

Patients’ most common presenting symptoms were cough (73.2%), fever (72.8%), and shortness of breath (63.1%) (Table 2). Dyspnea as a presenting symptom was significantly more common in those who progressed to the ICU, while non-ICU patients had the highest rates of nausea and vomiting. Notable lab findings on presentation include progressively higher inflammatory markers (C-reactive protein, erythrocyte sedimentation rate, ferritin, D-dimer, lactate dehydrogenase) for patients who would ultimately require ICU level of care as compared to hospitalized non-ICU patients and ED-only patients (Supplemental Table 3).

**Table 2.** Presenting Symptoms. The presenting symptoms of COVID-19 positive patients receiving care at NYP/CUIMC are detailed above in order of overall prevalence. All categorical data is reported as a frequency and a column percentage [95% confidence interval].

|  | <u>Highest Level of Care</u> |  |  |  |
| --- | --- | --- | --- | --- |
|  | **ED**<br>**(n = 150)** | **Hospitalized (non-ICU)**<br>**(n = 614)** | **ICU**<br>**(n = 236)** | **Overall**<br>**(N = 1000)** |
| Cough | 116<br>77.3% [70.0 – 83.3] | 447<br>72.8% [69.1 – 76.2] | 169<br>71.6% [65.5 – 77] | 732<br>73.2% [70.4 – 75.9] |
| Fever | 110<br>73.3% [65.7 – 79.8] | 447<br>72.8% [69.1 – 76.2] | 171<br>72.5% [66.4 – 77.8] | 728<br>72.8% [70.0 – 75.5] |
| Dyspnea | 83<br>55.3% [47.3 – 63.1] | 375<br>61.1% [57.2 – 64.9] | 173<br>73.3% [67.3 – 78.5] | 631<br>63.1% [60.1 – 66] |
| Myalgias | 44<br>29.3% [22.6 – 37.1] | 178<br>29% [25.5 – 32.7] | 46<br>19.5% [14.9 – 25.0] | 268<br>26.8% [24.1 – 29.6] |
| Diarrhea | 29<br>19.3% [13.8 – 26.4] | 157<br>25.6% [22.3 – 29.2] | 50<br>21.2% [16.5 – 26.8] | 236<br>23.6% [21.1 – 26.3] |
| Chills | 25<br>16.7% [11.6 – 23.4] | 122<br>19.9% [16.9 – 23.2] | 40<br>16.9% [12.7 – 22.3] | 187<br>18.7% [16.4 – 21.2] |
| Nausea/Vomiting | 15<br>10% [6.2 – 15.8] | 139<br>22.6% [19.5 – 26.1] | 24<br>10.2% [6.9 – 14.7] | 178<br>17.8% [15.6 – 20.3] |
| Headache | 21<br>14% [9.3 – 20.5] | 65<br>10.6% [8.4 – 13.3] | 15<br>6.4% [3.9 – 10.2] | 101<br>10.1% [8.4 – 12.1] |
| Sore Throat | 17<br>11.3% [7.2 – 17.4] | 48<br>7.8% [5.9 – 10.2] | 19<br>8.1% [5.2 – 12.2] | 84<br>8.4% [6.8 – 10.3] |
| Rhinorrhea | 16<br>10.7% [6.7 – 16.6] | 49<br>8% [6.1 – 10.4] | 18<br>7.6% [4.9 – 11.7] | 83<br>8.3% [6.7 – 10.2] |
| Sputum | 9<br>6% [3.2 – 11.0] | 57<br>9.3% [7.2 – 11.8] | 15<br>6.4% [3.9 – 10.2] | 81<br>8.1% [6.6 – 10.0] |
| Syncope | 6<br>4% [1.8 – 8.5] | 35<br>5.7% [4.1 – 7.8] | 7<br>3% [1.4 – 6.0] | 48<br>4.8% [3.6 – 6.3] |
| Conjunctival Congestion | 0<br>0% [0.0 – 2.5] | 2<br>0.3% [0.1 – 1.2] | 3<br>1.3% [0.4 – 3.7] | 5<br>0.5% [0.2 – 1.2] |

### Inpatient Hospital Course and Outcomes

Table 3 provides an overview of the hospital course of the inpatient cohort. Of the 1000 patients, 910 patients reached a primary endpoint as of April 30: 699 patients were discharged, 211 had died in the hospital, and 90 were still hospitalized. Of the 150 ED patients 85.3% of patients were discharged and 14.7% died prior to admission; of the 614 non-ICU patients, 14.0% died. There were 236 ICU patients; 93.2% were intubated at least once, 31.4% were extubated at least once, and 19.5% were discharged; 43.6% of ICU patients died in the hospital and 36.9% remain hospitalized. The majority of ICU patients (73.6%) required supplemental O2 within 3 hours of arriving at the ED and were on a nasal cannula (60.6%) or non-rebreather (73.7%) while less than 10% received high-flow nasal cannula or non-invasive positive pressure ventilation during their hospitalization (Supplemental Table 4). In our multivariate Cox models, age and BMI, along with pre-existing HIV or renal disease were statistically significantly associated with death, while gender and hypertension were associated with intubation and the composite outcome of intubation or death (Supplemental Table 5).

Overall, 64.9% of patients received over 48 hours of antibiotic therapy during their stay (most commonly azithromycin) and 63.9% received hydroxychloroquine (Table 3). Both treatments were more prevalent in ICU patients, with 94.9% on antibiotics and 89.8% on hydroxychloroquine; 94.1% of patients in the ICU received vasopressors at some point during their hospital course.

Across all patients with COVID-19, 33.9% developed acute kidney injury (AKI) and 13.8% required inpatient hemodialysis (Table 3). In the ICU, AKI and hemodialysis were even more common at 78.0% and 35.2%, respectively. Acute respiratory distress syndrome (ARDS) was diagnosed in 35.2% of all patients and 89.8% of ICU patients.

**Table 3.**
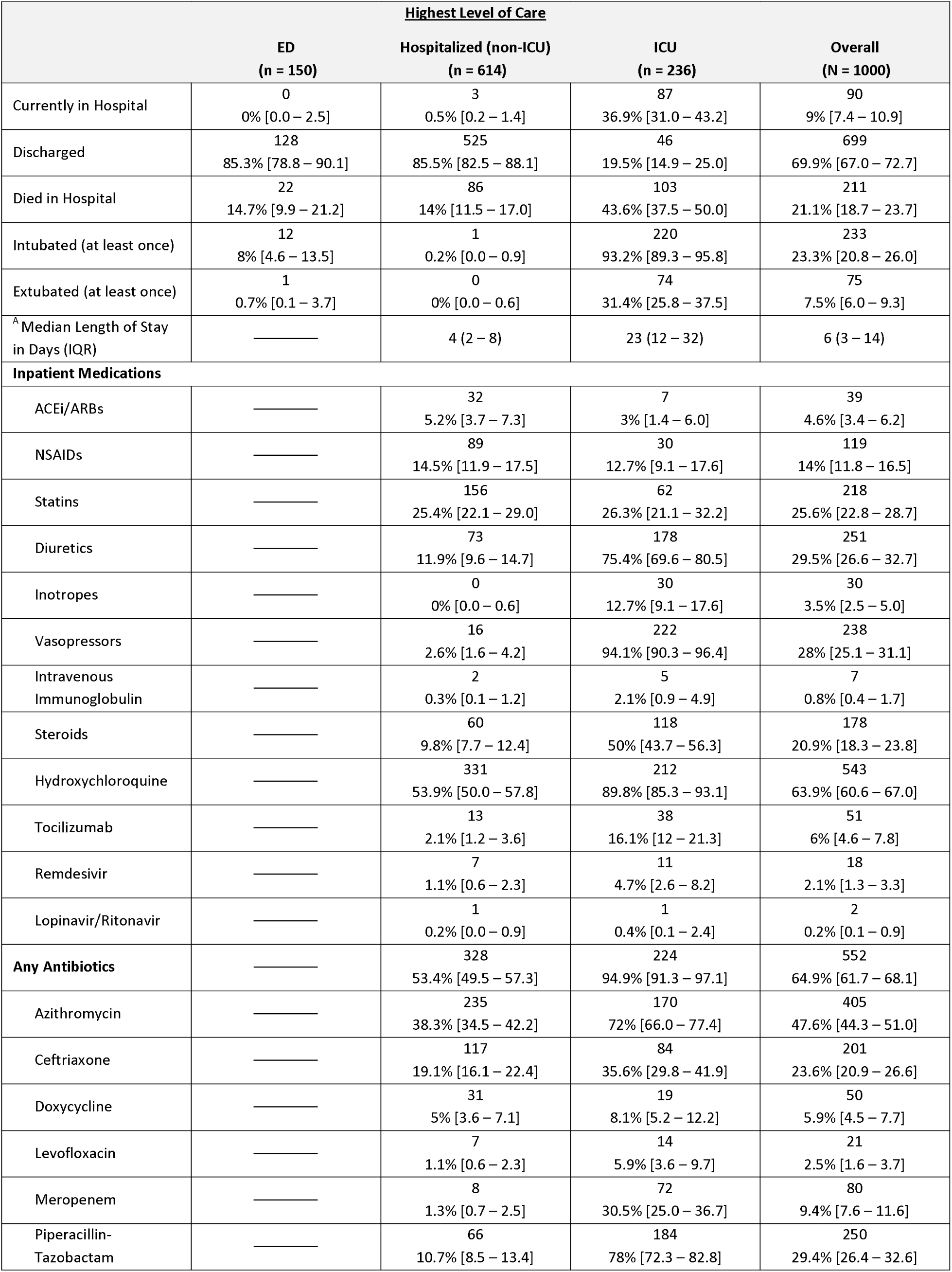
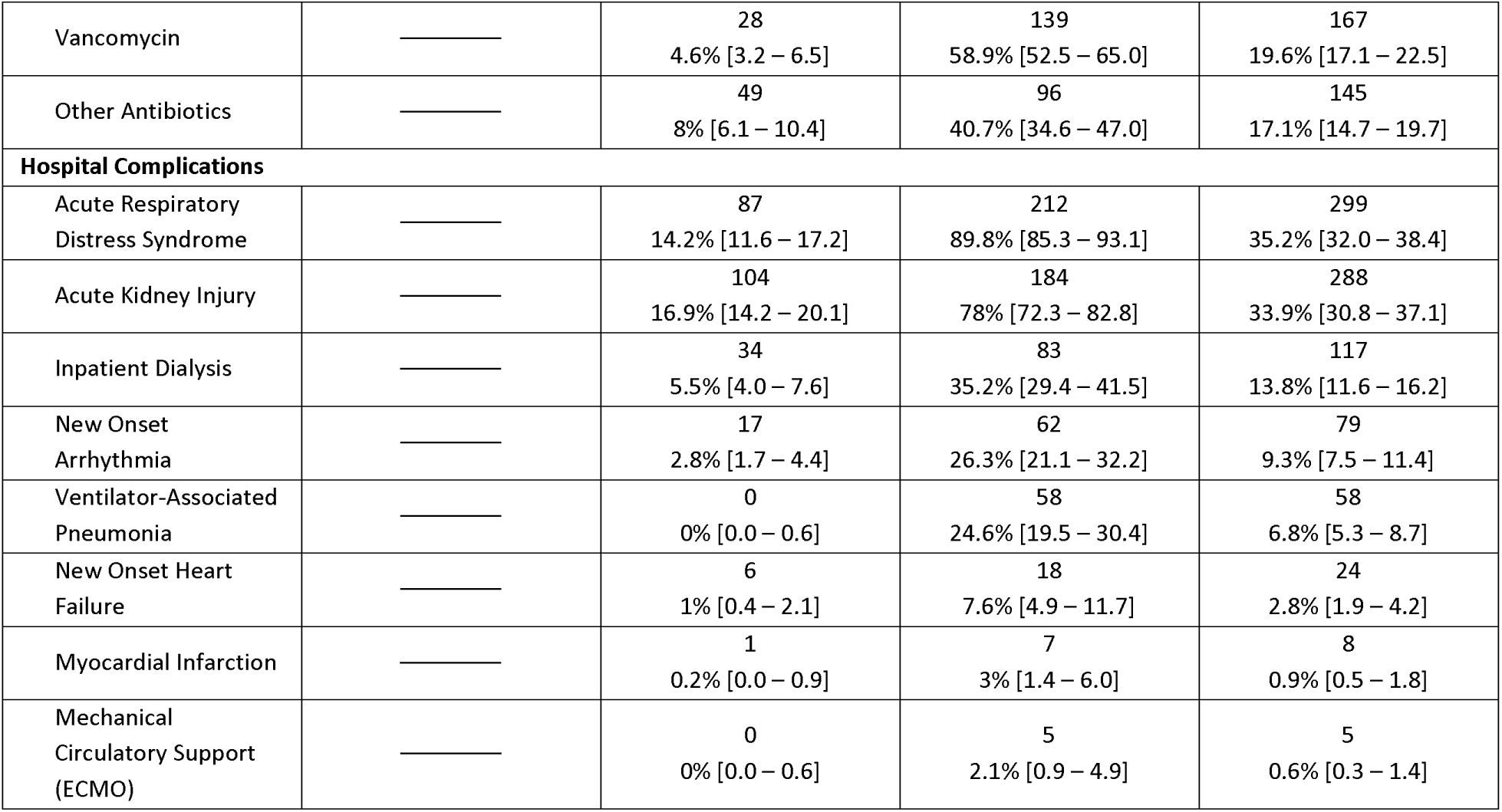
Inpatient Characteristics, Medications, and Complications. Patients receiving care at NYP/CUIMC ED were stratified according to the highest level of care received during their hospitalization. The intubated patient row includes all patients who were intubated at least once – they may be intubated, extubated, re-intubated, or dead. Extubated patients include only those who were successfully extubated, including patients who died later in their hospital course. It is important to note that 90 patients have not reached the end of their hospital course and that their charts continue to be reviewed. All categorical data is reported as a frequency and a column percentage [95% confidence interval]. All continuous data is reported as median (interquartile range). These are the outcomes reviewed as of April 30, 2020. ACEi, Angiotensin converting enzyme inhibitor; ARB, angiotensin receptor blocker; NSAIDs, nonsteroidal anti-inflammatory drugs. ^A^ Median length of stay (LOS) is calculated as days from admission to either discharge, death, or last chart review.

### Time Course of Intubated Patients

The time from the first reported symptoms to initial intubation (for the 136 intubated patients with exact date of first symptom recorded) appears bimodal (p = 0.004 for multimodality), with modes at 3–4 days and at 9 days post symptom onset (Figure 1). For patients who ultimately required mechanical ventilation, 95.6% were first intubated within 14 days after symptom onset. Additionally, 71.6% were intubated within the first three days after ED arrival (Supplemental Figure 1).

**Figure 1.**
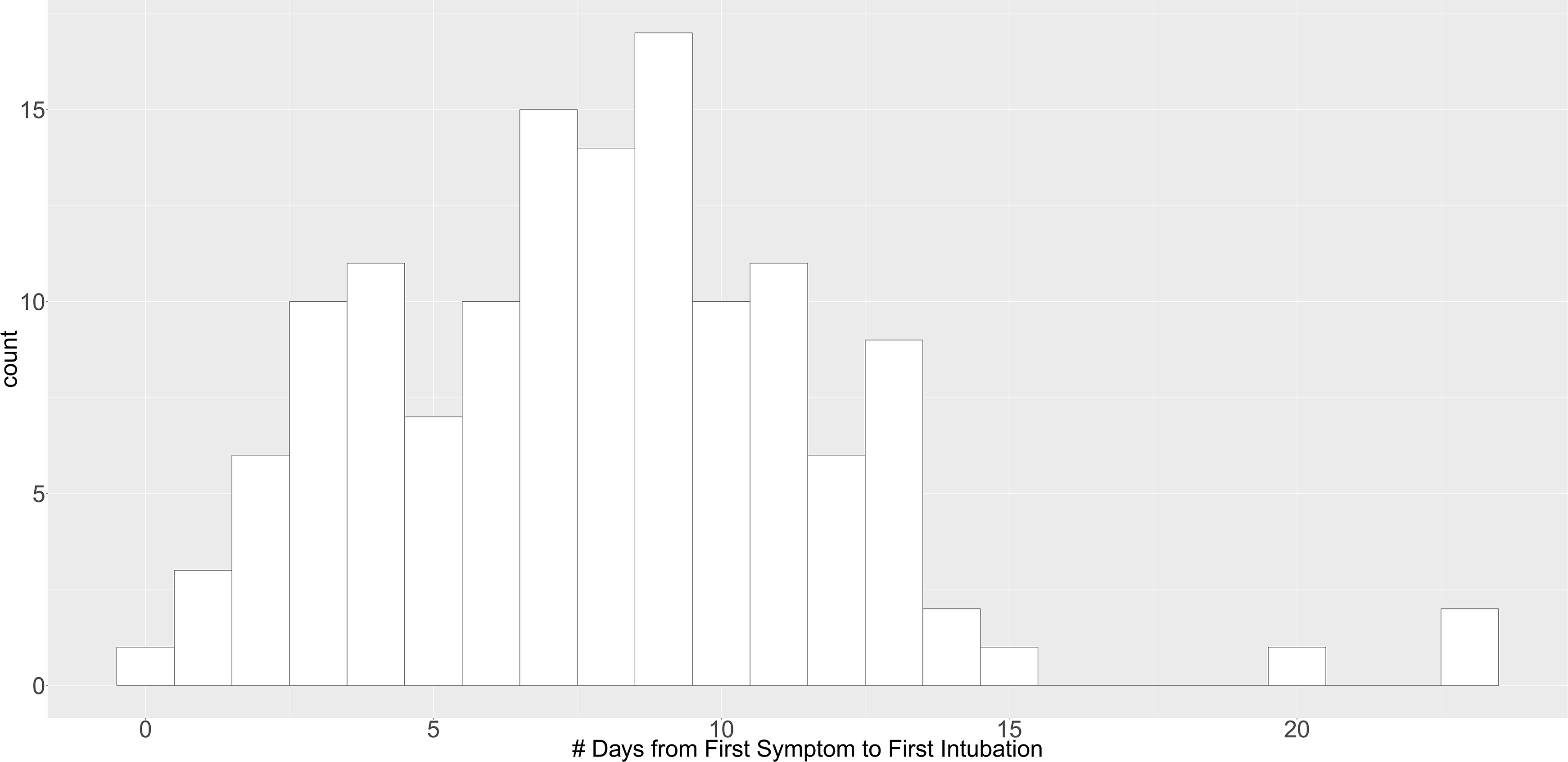
Distribution of Time from First Symptom to Intubation. 136 patients had a specific recorded date for first symptom onset. These patients are visualized in this figure. Days from first symptom to first intubation follows a bimodal distribution (p = 0.0088 for Hartigan’s Dip Test^18^), with modes at 3–4 days and 9 days. 4.4% of patients are intubated for the first time more than 14 days after the onset of symptoms.

Figure 2 displays the hospital timeline for each intubated patient, starting from ED presentation, and stratified by clinical status (death, discharge, or currently hospitalized). For the 233 intubated patients, 32.2 were extubated at least once, 47.6% died in the hospital, 15.5%, 5.7% were discharged from the hospital, and 36.9% were still hospitalized. Median (IQR) time of invasive mechanical ventilation (for first intubation) was 6.0 (2.0–13.0) days in patients who died, 9.0 (6.5–12.0) days in patients who were discharged, and 28.5 (22.5–31.75) days in patients who were still hospitalized.

**Figure 2.**
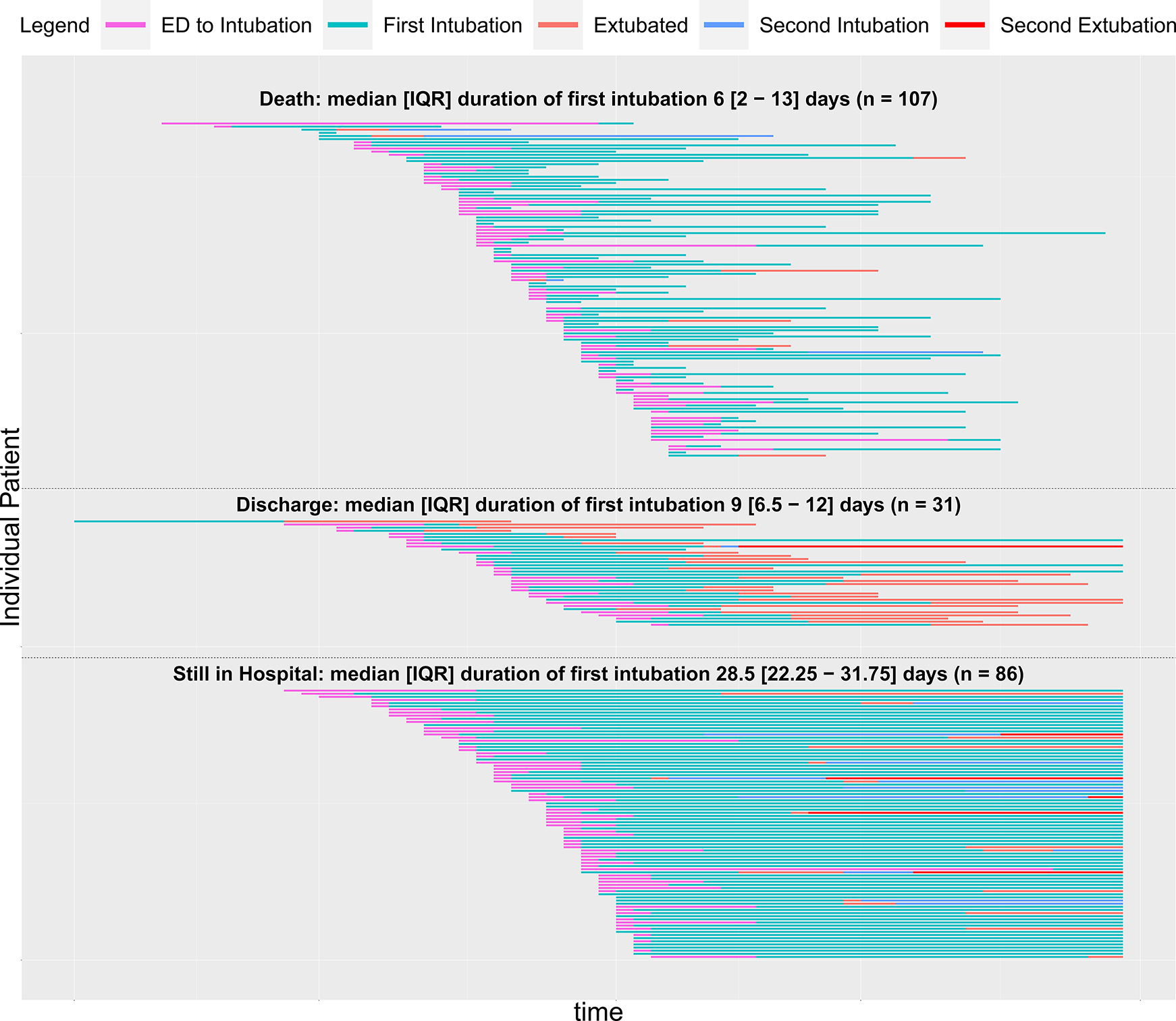
Timeline of Intubated Patients. The time course for COVID-19 positive patients intubated with exact times of intubation documented in the EHR (n=227^A^) at CUIMC is shown above. Patients are separated based on their current endpoints, separated into death, discharge, or currently hospitalized. 86 patients^8^ are still hospitalized, 31 have been discharged, 107 have died, and 74 have been extubated. ^A^7 patients acquired COVID-19 after intubation and are excluded from this plot. ^B^Data collection remains ongoing for currently hospitalized patients.

## DISCUSSION

In our characterization of the first 1000 consecutive patients with COVID-19 who received ED or inpatient care at NYP/CUIMC, we found a bimodal distribution for time to intubation from symptom onset. Our cohort had high baseline comorbidities and developed high rates of AKI and inpatient dialysis, along with prolonged intubation time and lengths of stay. Through manually abstracted data, this retrospective study provides an in-depth description of patients with COVID-19 at a more granular level than prior literature, including a previously undescribed bimodal distribution for time to intubation, which may suggest a biphasic nature to the COVID-19 disease process. We hope a better understanding of our patient population, baseline characteristics, inpatient course, and clinical outcomes can provide valuable guidance to clinicians who are working in a time of unparalleled volume and uncertainty.

As compared with prior literature, we found higher rates of renal complications in our patient sample. Previous studies from China reported 15% of all COVID-19 patients developed AKI,^11^ while a case series in Seattle found 19.1% developed AKI.^5^ However, we found 33.9% of all COVID-19 patients and 78.0% of ICU patients developed AKI, a striking increase compared to previous reports. Concomitantly, 13.8% of all patients and 35.2% of ICU patients required inpatient hemodialysis, leading to a scarcity of equipment needed for dialysis and continuous renal replacement therapy (CRRT). Similar experiences with slightly lower rates of AKI and CRRT have been reported in other NYC hospitals.^6^ This has resulted in the shared allocation of dialysis machines across patients, including the ICU setting. There may be multiple explanations for these differences. As part of treating patients with ARDS, providers often limit use of intravenous fluids, and this lung-protective fluid management strategy may have incidentally led to higher rates of AKI. Alternatively, there may be inherent renal toxicity associated with the pathophysiology of COVID-19 viral infection, given that the rates of AKI even in non-ICU patients or those without ARDS remain high. These rates may also be relatively higher than previous literature due to the relatively high acuity and increased comorbidities of our patient population.

Relative to previous cohorts, our patients have a higher average BMI, greater prevalence of hypertension, diabetes, and COPD, while fewer patients have no major comorbidity (8.2% vs. 32% and 52%) compared to those characterized in Italy and China.^10–13^ During the study period, NYC encouraged mildly symptomatic patients to stay home, and NYP/CUIMC implemented triaging practices (including cough/cold/fever clinics, initial evaluation in tents outside of ED and telemedicine follow-up) to manage patients without severe dyspnea at home. Thus, the patients who tested positive at NYP/CUIMC likely represented a higher acuity subset of symptomatic cases. Despite this, patients with COVID-19 in this sample have thus far had similar mortality to those presented in the epicenters of other countries. Across all levels of care, 21.1% of patients died, which is similar to NYU (18.5%)^15^ and lies in between estimates from China (1.4%, 28%).^11^ ^12^ ^21^ Our current mortality rate of ICU patients is 43.6%, while prior reports have suggested highly variable mortality rates in Italy (26%), China (38%, 78%), and Seattle (50%, 67%).^5^ ^7^ ^10^ ^11^ ^13^ However, since 36.9% of ICU patients remain hospitalized, the mortality will likely continue to rise.

The characterizations of prolonged intubation time and a bimodal distribution in time to intubation from symptom onset may help clinicians identify when patients are at high risk and anticipate disease progression. Of the 233 patients intubated at least once, 95.6% were intubated within the first 14 days of symptom onset, with bimodal peaks at 3-4 days and 9 days from symptom onset. Patients often undergo rapid respiratory decompensation, leading to increased clinician uncertainty. These findings may encourage plans for continued monitoring and vigilance despite clinical stability or improvement if patients are between the 3-4 and 9-day peaks. On the other hand, it may reassure providers to de-escalate or discharge when patients are on a stable or improving trajectory after 14 days of symptoms, thereby optimizing hospital beds and resource utilization. This pattern may be due to the underlying pathophysiology, different response groups or phenotypes of patients who develop critical illness at different times or changing practice patterns. However, as seen in Supplemental Figure 2, this distribution has not clearly varied over the course of this study, suggesting that practice patterns over time are less likely to be the primary factor behind this finding. In previous reports from China, patients tended to develop ARDS around day 12.^11^ Another paper from Italy hypothesized that older patients with COVID-19 tend to become dyspneic 5-7 days after symptom onset, while younger patients tend to develop dyspnea later.^8^ Further work is necessary to understand the mechanisms driving this distribution of intubation times, as it could dictate the timing of interventions and treatments.

Length of stay (LOS) and total time on mechanical ventilation remains high for these patients, with significant implications for post-recovery needs and sequelae. To date, our total cohort had a median LOS of 6 days, which increases to 23 days for ICU patients. The overall LOS was comparable to two cohorts in China with median LOS of 11 and 12 days respectively, but LOS for ICU patients is strikingly longer, with the former Chinese study reporting an ICU LOS of only 8 days.^11^ ^12^ In addition, our ICU median LOS will continue to rise, given that 36.9% of ICU patients were still hospitalized at last review. Median time on mechanical ventilation for our currently hospitalized patients was over 28 days and rising, which dramatically exceeds the LOS for the entire hospitalization of most patients in China. While the overall hospital course is comparable to prior influenza cohorts, the LOS of critically ill patients exceeds those of influenza patients who have reported median intubation durations of 10-12 days.^21^ ^22^ Understanding and anticipating this prolonged intubation course may help provide guidance on resource utilization and hospital capacity. Planning for post-ICU care for these patients will also be critically important, as lengthy intubations and hospital courses have profound implications for rehabilitation, critical illness neuropathy, discharge planning, physical therapy,^23–25^ increased home needs during a time of social distancing, and potential difficulty in returning to baseline functional status.

As the COVID-19 pandemic progresses, the characterization of these patients and outcomes may be more representative of the evolving clinical presentation and course that hospitals around the world may expect to see. These results can help guide the development of patient protocols (such as safe discharge guidelines and follow-up practices), inform emergency medical system responses, and drive the continued growth of telemedicine and remote monitoring.^26^ While an understanding of our experience may be helpful to hospitals and healthcare workers as they prepare to triage patients, we recognize that COVID-19 patients who require admission will have a high morbidity and mortality, and a high percentage will require ICU beds, ventilators, or dialysis. These sobering facts should motivate efforts to further investigate a potential biphasic disease course suggested by the distribution of intubations, model the resource needs across hospitals and countries based on these rising rates of complications, and continue to develop interventions to change the course of the disease.

## STUDY LIMITATIONS

This study has several limitations. First, data collection is limited to what is documented in the EHR. There may be errors in both patient recall and clinician documentation. Second, accuracy of data is limited by the accuracy of the data abstraction itself. We attempt to mitigate this potential error with both manual QC and by implementing a series of checks in the data after export from the REDCap database. Third, not all patients included in this paper have completed their hospital course and may have evolving outcomes or levels of care, although we now have minimum follow-up of 24 days. Data were collected from a single, urban academic medical center and may not be generalizable to all other regions. Lastly, multivariate modeling on this population may be limited by residual confounding and bias. However, the urgency for data to inform clinicians motivates us to provide this snapshot of patients at the point of last data abstraction on April 30, 2020. We deliberately focused on characterizing the data in this paper to provide descriptive statistics and figures rather than hypothesis driven statistical inference.

## CONCLUSION

Hospitalized patients with COVID-19 in NYC may be higher-risk at baseline with more comorbidities and develop more complications, as compared to previously published U.S. and international cohorts. These patients face significant morbidity and mortality, with high rates of AKI, dialysis, prolonged intubations, and a bimodal distribution of time to intubation from symptom onset. This characterization of COVID-19 patients in NYC may provide anticipatory guidance as the pandemic continues around the world.

What is already known on this topic
- Coronavirus disease 2019 (COVID-19) is a global pandemic, with New York City (NYC) as a new epicenter of the disease.
- The high burden of disease has quickly exceeded the standard capacity of hospital systems and raised concerns regarding optimal clinical management, safe maximization of hospital throughput, and resource allocation.
- Front-line healthcare providers have limited data to help anticipate the clinical course of these patients and how they compare to previous international cohorts.

What this study adds
- Patients with COVID-19 requiring mechanical ventilation had a bimodal distribution in time to intubation from symptom onset, with the vast majority (95.6%) first intubated within 14 days.
- Hospitalized patients, specifically those in ICUs, had more comorbidities compared to previous international cohorts, along with prolonged intubations and higher rates of AKI and dialysis.
- These findings may help inform front-line providers and provide anticipatory guidance for the international community during this pandemic.

## Data Availability

Requests for the statistical code and dataset can be made to the corresponding author.

## Acknowledgements/Statements

The authors wish to acknowledge the dedication, commitment and sacrifice of the staff, providers and personnel at our institutions through the local COVID crisis and express our profound sadness about the suffering and loss of our patients, their families and our community.

## Author Contributions

MGA, SLB, CLS, JRT contributed equally and share first authorship.

Study conception and design: MGA, SLB, CLS, JRT, GH, RC, KN, PG, MMS, JJC, FMB, LAC, KAE, ZPG, NH, SH, JHD, LEK, HL, AL, RL, ACS.

Acquisition, analysis, or interpretation of data: MGA, SLB, CLS, JRT, GH, RC, KHC, KN, PAA, FMB, RB, MFC, MIC, LAC, JHD, LBD, SAD, KAE, ZPG, DTG, AAH, NH, SH, LEK, HL, EL, AL, RL, AYS, ACS, ZES, CPS, YX, HWZ. Drafting of the manuscript: MGA, SLB, CLS, JRT

Critical revision of the manuscript for important intellectual content: MGA, SLB, CLS, JRT, GH, RC, MRB, BPC, NG, PG, AMM, AAP, MSR, NWS, SS, MDS, RGB, KHC, JJC, MMS, JEZ.

Statistical analysis: MGA, SLB, CLS, JRT.

Administrative, technical, or material support: MGA, SLB, CLS, JRT, SS, GH, RC, KN.

Study supervision: GH, RC, KN.

GH and RC are the guarantors of the study

### Additional Contributions

The authors would like to acknowledge the Weill Cornell Medicine COVID-19 Registry Team, which developed the chart abstraction tool used in this study and assisted with training of the VP&S medical students in the chart abstraction process.

### Funding/Support

This study received no specific funding or grant from any agency in the public, commercial, or not-for-profit sectors. RGB is supported by grant R01HL077612 and R01HL093081 from the U.S. National Institute of Health. KHC is supported by grant T32HL007854 from U.S. National Institutes of Health/National Heart, Lung, and Blood Institute. JJC is supported by grants from the U.S. National Institute of Health/National Center for Advancing Translational Science. GH is supported by grant R01 LM006910 from the U.S. National Institute of Health/National Library of Medicine. RC is supported by grant T15 LM007079 from the U.S. National Institute of Health/National Library of Medicine.

No funding organization or sponsor was involved in the design and conduct of the study; collection, management, analysis, and interpretation of the data; preparation, review, or approval of the manuscript; and decision to submit the manuscript for publication.

### Ethics/IRB Approval

This study was approved by the Columbia University Institutional Review Board under protocol AAAS9834.

### Data Sharing

Requests for the statistical code and dataset can be made to the corresponding author.

### Exclusive License/Copyright

The Corresponding Author has the right to grant on behalf of all authors and does grant on behalf of all authors, a worldwide license (http://www.bmi.com/sites/default/files/BMJ%20Author%20Licence%20March%202013.doc) to the Publishers and its licensees in perpetuity, in all forms, formats and media (whether known now or created in the future), to i) publish, reproduce, distribute, display and store the Contribution, ii) translate the Contribution into other languages, create adaptations, reprints, include within collections and create summaries, extracts and/or, abstracts of the Contribution and convert or allow conversion into any format including without limitation audio, iii) create any other derivative work(s) based in whole or part on the on the Contribution, iv) to exploit all subsidiary rights to exploit all subsidiary rights that currently exist or as may exist in the future in the Contribution, v) the inclusion of electronic links from the Contribution to third party material where-ever it may be located; and, vi) license any third party to do any or all of the above. All research articles will be made available on an open access basis (with authors being asked to pay an open access fee—see http://www.bmi.com/about-bmi/resources-authors/forms-policies-and-checklists/copyright-open-access-and-permission-reuse). The terms of such open access shall be governed by a <u>Creative Commons</u> license—details as to which Creative Commons license will apply to the research article are set out in our worldwide license referred to above.

### Data dissemination statement

Dissemination to study participants and/or patient organizations is not applicable.

### Transparency declaration

RC affirms that the manuscript is an honest, accurate, and transparent account of the study being reported. No important aspects of the study have been omitted, and all discrepancies from the study as planned have been explained.

### Patient and Public Involvement Statement

No patients were involved in setting the research question or the outcome measures, nor were they involved in developing plans for recruitment, design, or implementation of the study. No patients were asked to advise on interpretation or writing up of results. While there are no plans to disseminate the results of the research to study participants or the relevant patient community, the preliminary results are publicly available on medRxiv.

